# Association between Alzheimer’s disease and COVID-19: A bidirectional Mendelian randomization

**DOI:** 10.1101/2020.07.27.20163212

**Authors:** Di Liu, Xiaoyu Zhang, Weijie Cao, Jie Zhang, Manshu Song, Weijia Xing, Wei Wang, Qun Meng, Youxin Wang

## Abstract

**Background:** In observational studies, Alzheimer’s disease (AD) has been associated with an increased risk of Coronavirus disease 2019 (COVID-19), and the prognosis of COVID-19 can affect nervous systems. However, the causality between these conditions remains to be determined.

**Methods:** This study sought to investigate the bidirectional causal relations of AD with COVID-19 using two-sample Mendelian randomization (MR) analysis.

**Results:** We found that genetically predicted AD was significantly associated with higher risk of severe COVID-19 (odds ratio [OR], 3.329; 95% confidence interval [CI], 1.139-9.725; *P*=0.028). It’s interesting that genetically predicted severe COVID-19 was also significantly associated with higher risk of AD (OR, 1.004; 95% CI, 1.001-1.007; *P*=0.018). In addition, the two strong genetic variants associated with severe COVID-19 was associated with higher AD risk (OR, 1.018; 95% CI, 1.003-1.034; *P*=0.018). There is no evidence to support that genetically predicted AD was significantly associated with COVID-19 susceptibility, and vice versa. No obvious pleiotropy bias and heterogeneity were observed.

**Conclusion:** Overall, AD may causally affect severe COVID-19, and vice versa, performing bidirectional regulation through independent biological pathways.

## Introduction

Coronavirus disease (COVID-19), which is caused by the severe acute respiratory syndrome coronavirus 2 (SARS-COV2) and represents the causative agent of a potentially fatal disease, rapidly developing into a global pandemic^1^.The prognosis of COVID-19 can cause impaired vision, loss of smell, and affect reproductive and nervous systems^2 3^, that is of great global public health concern. A wide range of studies focused on susceptible populations for COVID-19. In the susceptible population of the elderly, specific risk factors such as dementia have been found to be a risk factor (odds ratio [OR] = 3.50, 95% CI: 1.93 to 6.34) for COVID-19 severity in the UK Biobank (UKB)^4^. However, findings from epidemiological studies are prone to unmeasured confounding, and reverse causation.

Mendelian randomization (MR)has becoming a popular method by which genetic variants robustly associated with exposures can be used as instrumental variables to infer causality from routinely conducted observational studies^5^. Summary Mendelian randomization (SMR) is an excellent strategy to evaluate the causality using summary statistics from GWAS data^6^. Therefore, we performed bidirectional MR analyses for causal inference between Alzheimer’s disease (AD), the main type of dementia, and COVID-19 using summary statistics GWAS results of AD and COVID-19. Understanding the bidirectional relations between AD and COVID-19 is of significant public health importance about disease prevention and management of complications.

## Methods

### Data sources

#### Genetic association datasets for COVID-19

The summarized data was obtained from the most recent version of GWAS analyses from the COVID-19 host genetics initiative from UK Biobank individuals, which released on July 1, 2020 (https://www.covid19hg.org/results/)^7^. Summarized data on COVID-19 included 3523 patients and 36634 control participants, and summarized data on severe COVID-19 included 536 patients and 329391 control participants. In addition, we performed this analysis with another summarized data from the GWAS of severe COVID-19 with respiratory failure reported by Severe Covid-19 GWAS Group^8^ including 835 patients and 1255 control participants from Italy as well as 775 patients and 950 control participants from Spain for supplementary analysis.

#### Genetic association datasets for Alzheimer’s disease

Summarized data on AD were obtained from the International Genomics of Alzheimer Project with 17008 Alzheimer’s disease cases and 37154 controls (https://www.pasteur-lille.fr)^9^. All GWAS summary statistic data were based on European ancestry populations.

### Instrumental variables for AD and COVID-19

For the bidirectional MR analysis, we first applied forward MR analysis to assess the effect of predisposition to AD on the phenotypes of COVID-19 or severe COVID-19 by using genetic variants robustly associated with AD. Then, we performed reverse MR using genetic variants robustly associated with the phenotypes of COVID-19 or severe COVID-19 as genetic instruments to investigate their effect on AD.

Single nucleotide polymorphisms (SNPs) that achieved significance (*P*<5×10^−8^) for the AD were selected as instrumental variables (IVs). SNPs were chosen as IVs for COVID-19 or severe COVID-19 at a significance threshold (*P*<1×10^−5^) from the COVID-19 host genetics initiative, because we didn’t filter out SNPs as IVs for COVID-19 or severe COVID-19 at a significance threshold (*P*<5×10^−8^). In addition, we chose two SNPs (rs11385942 and rs657152) as IVs for severe COVID-19 based on the GWAS of severe COVID-19 with respiratory failure reported by Severe Covid-19 GWAS Group based on the thresholds for genome-wide significance (*P*<5×10^− 8^) for supplementary analysis.

We only retained independent variants from each other (Linkage disequilibrium [LD], r^2^ < 0.001). When we encountered genetic variants in LD, the SNP with the lowest *p*-value was selected. The LD proxies were defined using 1000 genomes European samples. The same method was applied to filter instrumental variables in reverse MR analysis.

### MR analysis

In the main analyses, inverse-variance weighted (IVW) method was used to estimate the overall causal association of AD on COVID-19 and severe COVID-19 susceptibility. We additionally conducted sensitivity analyses using the weighted median, penalised weighted median, and MR-Egger regression to account for potential violations of valid instrumental variable assumptions. The MR-Egger analysis was performed to valuate pleiotropy based on the intercept. We conducted heterogeneity test in MR analyses using IVWQ test and a leave one out analysis in which we omitted one genetic variant to investigate the influence of pleiotropic genetic variants. Results are presented as odds ratios with their 95% confidence intervals (95% CIs) of outcomes per genetically predicted increase in each exposure factor. All data analyses were performed by “twosampleMR” package in R (R version 3.6.3, R Core Team, 2017).

In terms of various estimates for different measures, we chose the result of main MR method as following rules:

1. If no directional pleiotropy in MR estimates (Q statistic: *P* value > 0.05, MR-Egger intercept: *P* value > 0.05), we reported the results of IVW method.
2. If directional pleiotropy was detected (MR-Egger intercept: *P* value < 0.05) and *P* value > 0.05 for the Q test, we reported the results of MR-Egger method.
3. If directional pleiotropy was detected (MR-Egger intercept: *P* value < 0.05) and *P* value < 0.05 for the Q test, we reported the results of weighted median method.

The causal relationships between AD and COVID-19 were delineated into four potential categories, which were observed by bidirectional MR. **Figure 1** provided an overview of the bidirectional MR study to investigate these explanations. If the *P* value was less than 0.05 only in forward MR, genetic variants determined AD was associated with higher COVID-19 risk (Explanation 1). To robustly test Explanation 2, we performed the reverse MR analysis, evaluating whether COVID-19 affected AD. A *P* value greater than 0.05 indicated that Explanation 2 was unlikely. There are bidirectionally causal associations between AD and COVID-19, if both *P* values were less than 0.05 in forward and reverse MR. We identified the results to evaluate Explanation 3, that is, the finding is based on that the selected SNPs as IVs for forward and reverse MR analysis were not overlapped between AD and COVID-19. If the *P* value was greater than 0.05 in forward and reverse MR, it was interpreted as Explanation 4.

**Figure 1.**
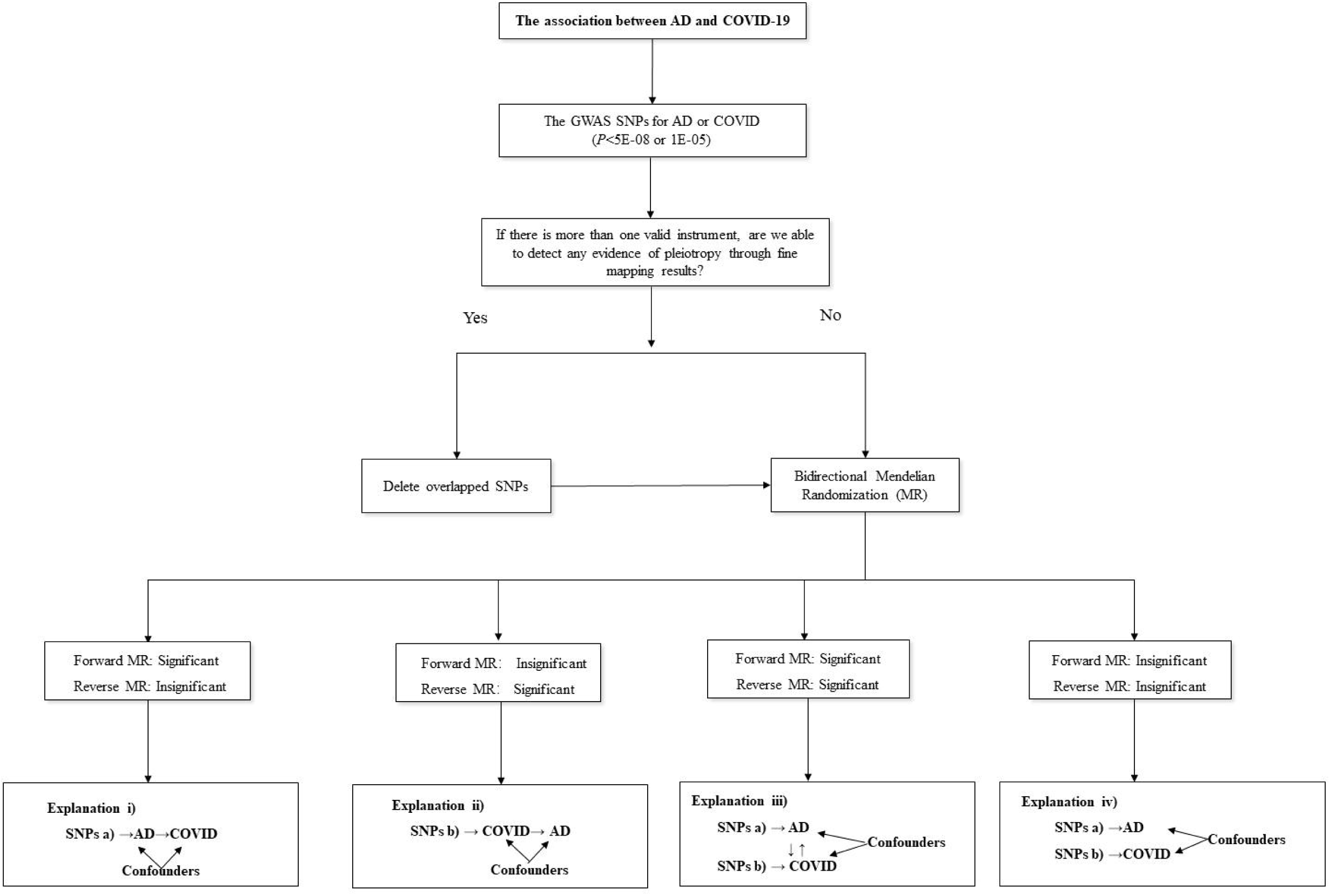
Analysis pipeline to evaluate the explanations for the observed associations between AD and COVID-19 AD: Alzheimer’s disease; COVID-19: Coronavirus disease 2019; GWAS: genome-wide association study; MR: mendelian randomization; SNP, single-nucleotide polymorphism

The annotations of SNPs residing in ± 100 kb of the corresponding genes were obtained from annotation files of the Grch37/hg19. To functionally annotate putative SNPs, we conducted functional enrichment analysis for the identified genes using the functional annotation tool “Metascape”^10^ and the R package “clusterProfiler”. Gene symbols corresponding to putative genes (*P* < 0.05) were used as the input of the gene ontology (GO) and Kyoto Encyclopedia of Genes and Genomes enrichment analysis.

## Results

### Causal association of AD with COVID-19 via forward MR

In the AD→COVID-19 MR analysis, we used 20 independent SNPs as the instrument for AD. As shown in **Table 1**, IVW estimate showed there was no association between the genetically instrumented AD and COVID-19 risk (OR, 1.211; 95%CI, 0.775-1.895; *P*=0.401), without obvious pleiotropy bias (intercept=-0.006, *P*=0.673) and heterogeneity (*P*=0.091). Of note, there was association of the genetically instrumented AD with severe COVID-19 using 19 SNPs (OR, 3.329; 95% CI, 1.139–9.725, *P*=0.028), without directional pleiotropy (intercept=-0.028, *P*=0.318) and heterogeneity (*P*=0.335). The association of AD with COVID-19 and severe COVID-19 remained robust in the weighted median and penalised weighted median methods, and leave one out analyses (**Figure S1** and **Figure 2**).

**Table 1.** Causal association of AD with COVID-19 via forward MR.

| Phenotype | IVs for AD (SNPs) | OR (95% CI) | Beta (SE) | *P* |
| --- | --- | --- | --- | --- |
| COVID vs. Lab/self-reported negative |  |  |  |  |
| IVW | 20 | 1.211 (0.775-1.895) | 0.192 (0.228) | 0.401 |
| Weighted median | 20 | 1.145 (0.697-1.880) | 0.135 (0.253) | 0.593 |
| Penalised weighted median | 20 | 1.136 (0.704-1.831) | 0.127 (0.244) | 0.601 |
| MR_Egger | 20 | 1.340 (0.699-2.569) | 0.293 (0.332) | 0.389 |
| $\beta$ (intercept) | - | - | -0.006 (0.014) | 0.673 |
| Q statistic | - | - | - | 0.091 |
| Severe respiratory confirmed COVID vs. population |  |  |  |  |
| IVW | 19 | 3.329 (1.139-9.725) | 1.203 (0.547) | 0.028 |
| Weighted median | 19 | 4.691 (1.282-17.165) | 1.546 (0.662) | 0.019 |
| Penalised weighted median | 19 | 4.874 (1.353-17.558) | 1.584 (0.654) | 0.015 |
| MR_Egger | 19 | 5.405 (1.314-22.238) | 1.687 (0.722) | 0.032 |
| $\beta$ (intercept) | - | - | -0.028 (0.027) | 0.318 |
| Q statistic | - | - | - | 0.335 |

Beta is the estimated effect size.
AD: Alzheimer's disease; CI: confidence intervals; IVs: instrumental variables; IVW, inverse-variance weighted; MR mendelian randomization; OR: odds ratio; SE, standard error; SNP, single-nucleotide polymorphism.

**Figure 2.**
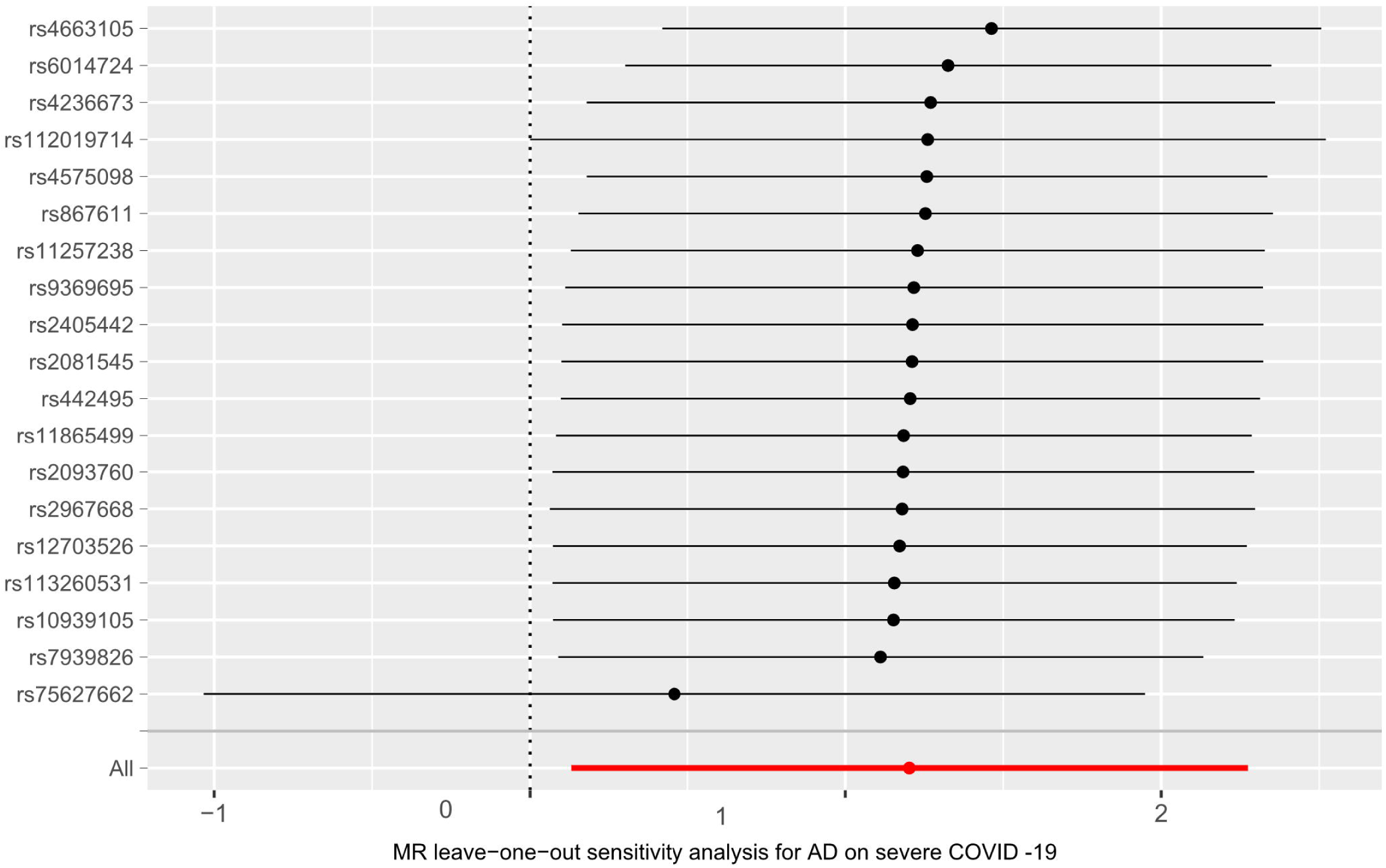
Leave-one-out analysis: each row represents a MR analysis of AD on severe COVID-19 using all instruments expect for the SNP listed on the y-axis. The point represents the odds ratio with that SNP removed and the line represents 95% confidence interval. AD: Alzheimer’s disease; COVID-19: Coronavirus disease 2019; MR: mendelian randomization; SNP, single-nucleotide polymorphism

### Causal association of COVID-19 with AD via reverse MR

As shown in Table 2, the reverse MR analysis showed no significant association of genetically instrumented COVID-19 with AD using 17-SNP instrument (OR, 0.999; 95% CI, 0.990-1.007; *P*=0.756). The association of COVID-19 with AD was robust in the weighted median and penalised weighted median methods, and leave one out analyses (**Figure S2**). Pleiotropy bias (*P* for MR-Egger intercept=0.263) and heterogeneity (*P*=0.738) were also not detected. It is worth mentioning that the genetically instrumented severe COVID-19 was causally associated with AD using 15-SNP instrument (OR, 1.004; 95% CI, 1.001–1.007; *P*=0.018). The causal effect of severe COVID-19 on AD was not robust using weighted median (OR, 1.004; 95% CI, 0.999-1.009; *P*=0.117), penalised weighted median (OR, 1.004; 95% CI,0.999-1.008; *P*=0.112) and MR-Egger regression (OR, 0.999; 95% CI,0.989-1.008; *P*=0.820), respectively. There was limited evidence of heterogeneity and horizontal pleiotropy based on the Q test (*P*=0.738) and MR-Egger intercept test (*P*=0.263). In terms of various estimates for different measures, we reported the results of IVW method. The association of severe COVID-19 with AD remained robust in the leave one out analyses (**Figure 3**). In addition, genetically predicted severe COVID-19 was significantly associated with higher risk of AD using the two strong genetic variants of rs11385942 (locates at 3p21.31) and rs657152 (9q34.2, ABO blood-group) as IVs for severe COVID-19 (OR, 1.018; 95% CI, 1.003-1.034; *P*=0.018).

**Table 2.** Causal association of COVID-19 with AD via reverse MR.

| Phenotype | IVs for COVID (SNPs) | OR (95% CI) | Beta (SE) | *P* |
| --- | --- | --- | --- | --- |
| COVID vs. Lab/self-reported negative |  |  |  |  |
| IVW | 17 | 0.999 (0.990-1.007) | -0.0013 (0.0043) | 0.756 |
| Weighted median | 17 | 1.0003 (0.989-1.011) | 0.0002 (0.0056) | 0.960 |
| Penalised weighted median | 17 | 1.0006 (0.989-1.011) | 0.0006 (0.0055) | 0.919 |
| MR_Egger | 17 | 1.0015 (0.977-1.026) | 0.0015 (0.0123) | 0.905 |
| $\beta$ (intercept) | - | - | -0.0007 (0.0028) | 0.891 |
| Q statistic | - | - | - | 0.421 |
| Severe respiratory confirmed COVID vs. population |  |  |  |  |
| IVW | 15 | 1.004 (1.0007-1.007) | 0.0042 (0.0018) | 0.018 |
| Weighted median | 15 | 1.004 (0.999-1.009) | 0.0038 (0.0024) | 0.117 |
| Penalised weighted median | 15 | 1.004 (0.999-1.008) | 0.0038 (0.0024) | 0.112 |
| MR_Egger | 15 | 0.999 (0.989-1.008) | -0.0011 (0.0049) | 0.820 |
| $\beta$ (intercept) | - | - | 0.0025 (0.0021) | 0.263 |
| Q statistic | - | - | - | 0.738 |
| Severe respiratory confirmed COVID vs. population* |  |  |  |  |
| IVW | 2 | 1.018 (1.003-1.034) | 0.018 (0.008) | 0.018 |

\*The two SNPs (rs11385942 and rs657152) were selected as IVs for severe COVID-19 based on the GWAS of severe COVID-19 with respiratory failure reported by Severe Covid-19 GWAS Group based on thresholds for genome-wide significance ( $P < 5 \times 10^{-8}$ ).
Beta is the estimated effect size.
AD: Alzheimer's disease; CI: confidence intervals; IVs: instrumental variables; IVW, inverse-variance weighted; MR mendelian randomization; OR: odds ratio; SE, standard error; SNP, single-nucleotide polymorphism

**Figure 3.**
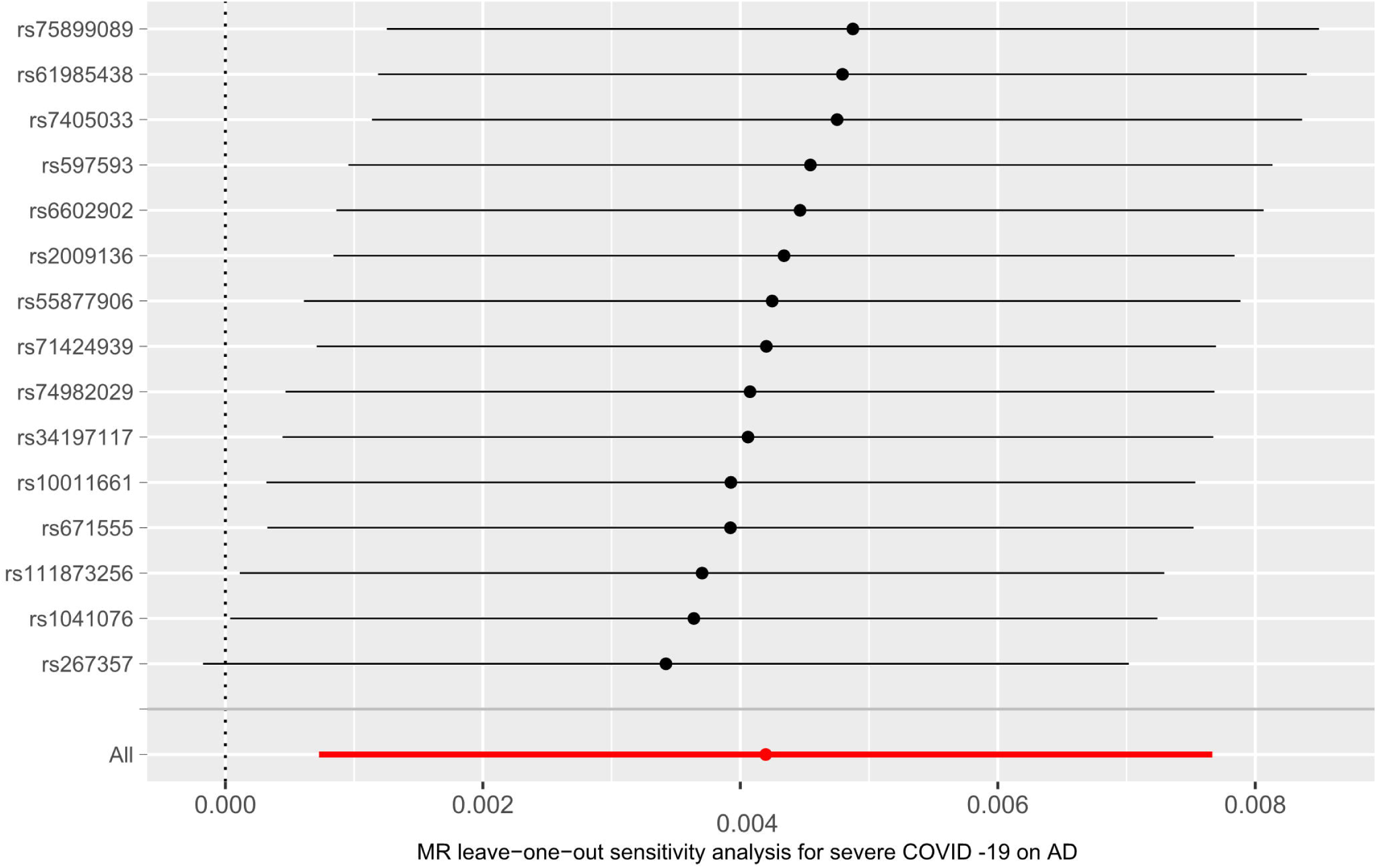
Leave-one-out analysis: each row represents a MR analysis of severe COVID-19 on AD using all instruments expect for the SNP listed on the y-axis. The point represents the odds ratio with that SNP removed and the line represents 95% confidence interval. AD: Alzheimer’s disease; COVID-19: Coronavirus disease 2019; MR: mendelian randomization; SNP, single-nucleotide polymorphism

### Functional bioinformatics

The SNPs as IVs for AD were annotated to 126 unique genes (**Table S1**). GO enrichment analysis of biological process, molecular function, and cellular component pathways showed that the identified genes were involved in 20 GO terms (**Figure 4**), such as negative regulation of amyloid precursor protein catabolic process, immunoglobulin mediated immune response, regulation of GTPase activity, cellular oxidant detoxification, regulation of GTPase activity. *APOE, ABCA7, BIN1, SORL1* and *CLU* were involved in the pathway of negative regulation of amyloid precursor protein catabolic process and amyloid beta formation. The SNPs as IVs for severe COVID-19 were annotated to 31 unique genes (**Table S2**). GO enrichment showed that the identified genes were involved in 3 GO terms (**Figure 5**), including regulation of chromosome organization, post-translational protein modification and positive regulation of GTPase activity. *DRD1* and *MCTP1* were involved in the pathway of regulation of neurotransmitter secretion and transport. It is important to note that the shared pathway of the regulation of GTPase activity might link AD and severe COVID-19.

**Figure 4.**
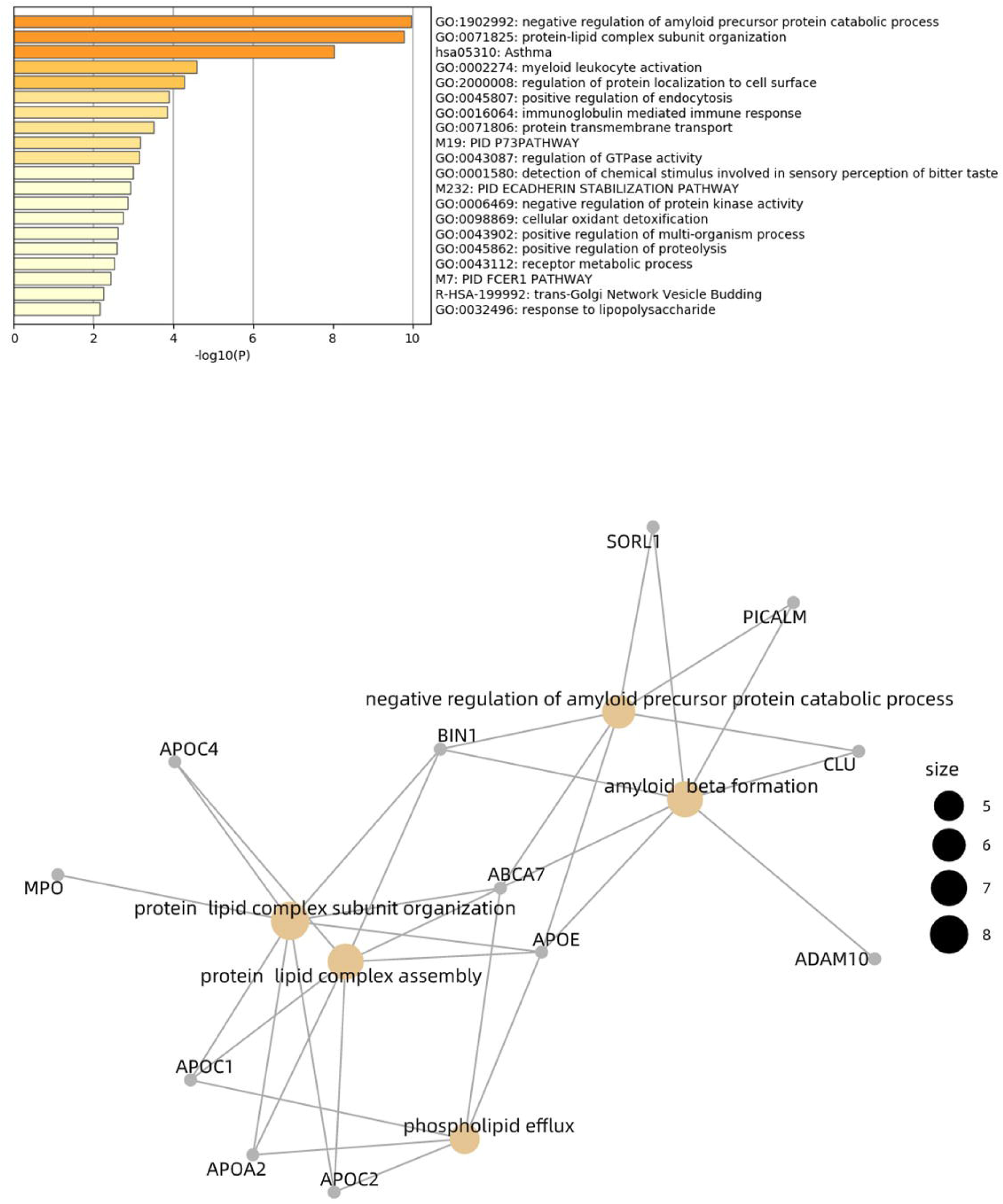
Functional enrichment analysis of the genes annotated by SNPs as IVs for AD. A) Enriched GO terms via the genes; B) The association between genes and the enriched GO clusters AD: Alzheimer’s disease; IVs: instrumental variables; GO, gene ontology; SNP, single-nucleotide polymorphism

**Figure 5.**
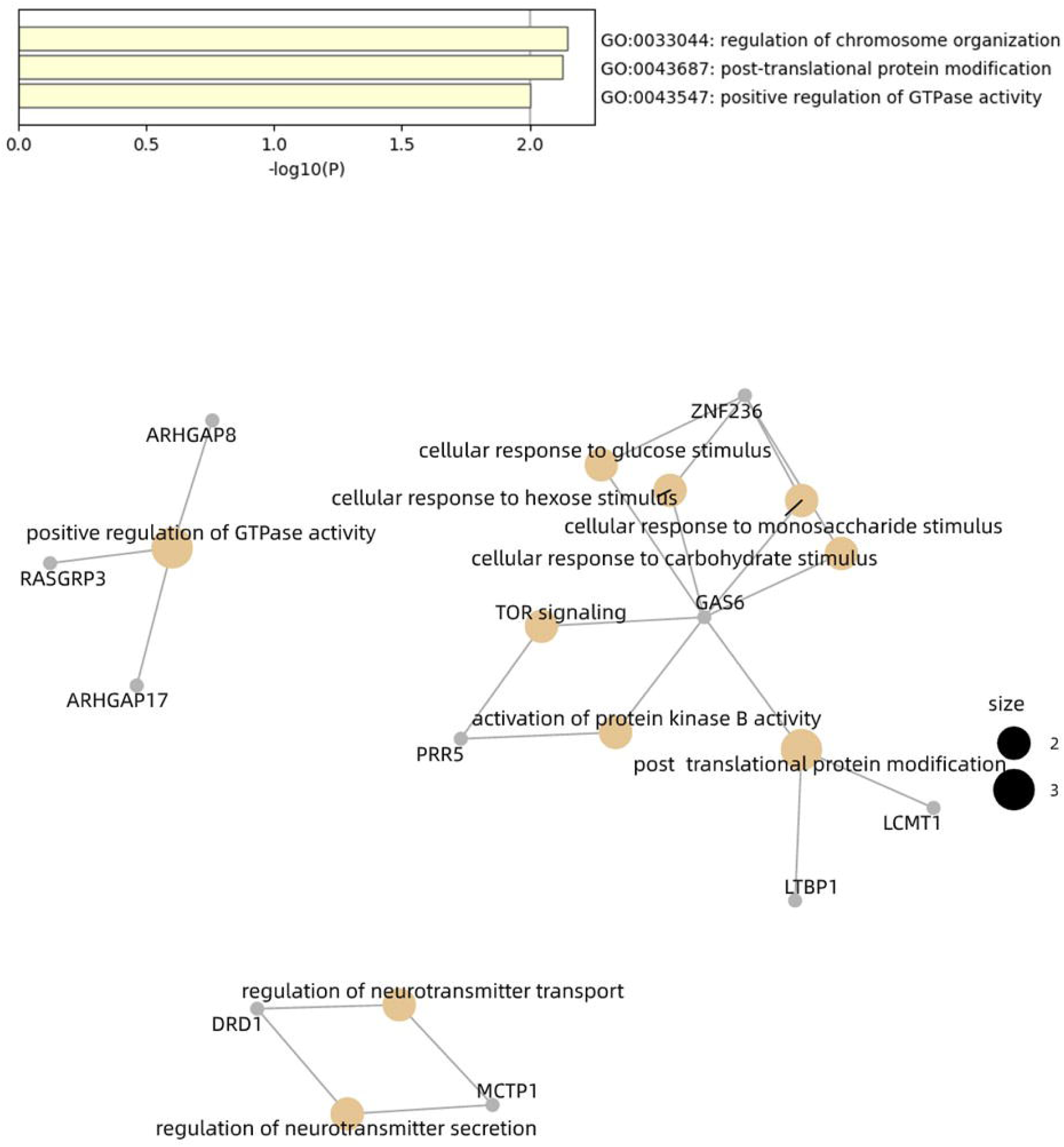
Functional enrichment analysis of the genes annotated by SNPs as IVs for severe COVID-19. A) Enriched GO terms via the genes; B) The association between genes and the enriched GO clusters COVID-19: Coronavirus disease 2019; IVs: instrumental variables; GO, gene ontology; SNP, single-nucleotide polymorphism

## Discussion

To our knowledge, this is the first study to investigate the causal relationship between AD and severeCOVID-19 using a bidirectional MR. Our study showed that AD may causally affect severe COVID-19, and vice versa, performing bidirectional regulation through independent biological pathways.

Our study found that AD was causally associated with the susceptibility of severe COVID-19, but not COVID-19, might indicating that AD patients infected with severe COVID-19 were more likely to be severe phenotype. The finding was consistent with the observational epidemiology study, which found that pre-existing dementia was associated with the risk of COVID-19 hospitalization and death increase ^4^. In addition, the study showed that *ApoE*ε4ε4 homozygotes were more likely to be COVID-19 positives (OR = 2.31, 95% CI: 1.65 to 3.24, *P* = 1.19E–6) compared to ε3ε3 homozygotes in the UKBiobank Community Cohort. The *ApoE*ε4ε4 genotype was associated with a 14-fold increase in risk of AD compared to the common ε3ε3 genotype in populations with European ancestries^11^. Therefore, AD patients with the *ApoE* ε4ε4 allele might increase risks of severe COVID-19 infection^12^. In addition, the genes annotated by genetic variants strongly associated with AD were involved in the pathway of immune response. That could explain the death of most severe COVID-19 patients due to the inflammatory storm ^13 14^.

This is the first study to identify the prognosis of COVID-19. We found that severe COVID-19 might cause AD. Notably, the COVID-19 cases endanger the elderly, especially those with cardiovascular illness including diabetes mellitus, hypertension, and coronary heart disease^15 16^. The increasing MR studies have found that coronary artery disease, higher body mass index, lifetime smoking and dyslipidemia were causally associated with the susceptibility of COVID-19^17-20^, which were also risk factors for AD^21 22^. The reasons underlying AD also may relate to the proinflammatory effect associated with severe COVID-19. An exaggerated inflammatory response appears a major driver of immunopathology in SARS-CoV-2 infected patients^23^, with benefits of targeting the acute inflammatory response in severe COVID-19 cases^24^. It has been reported that bi-directional communication existed between the central nervous system and the peripheral immune system^25^. Activated peripheral immune cells that enter the brain through the blood brain barrier (BBB) may modulate the pathogenesis of AD^26 27^. The genes *DRD1* and *MCTP1* annotated by genetic variants strongly associated with severe COVID-19 were involved in the pathway of regulation of neurotransmitter secretion and transport. SARS-COV2 might be a causative factor involved in the infectious etiology of AD. In addition, ABO blood type might play an important role in the development of COVID-19 and AD^8 28^. Taken together, the findings indicated that subjects infected with severe COVID-19 with respiratory failure might be of higher risk of AD after survival of COVID-19.

No overlapped genetic variants as IVs for AD and severe COVID-19 was overserved, indicating that AD and severe COVID-19 might be bidirectionally regulated by different pathogenic pathways. The shared pathway of the regulation of GTPase activity might underly the association between AD and severe COVID-19 ^29-31^, which needed further investigation to get more data support and further experimental study to verify.

The bidirectional MR analysis aims to explore positive and negative feedback loops resulting in regulation of the COVID -19 as a causal or a downstream effect on diseases. We should notice some limitations. First of all, stratified analyses or analyses adjusted for other covariates were not possible on the account of using available summary statistics datasets. The GWAS of Severe COVID-19 with respiratory failure and the COVID-19 host genetics initiative from UK Biobank individuals include small sample size, which may lead to small effect for the MR estimate and limit the IVs for COVID-19 for reverse MR analysis. The annotated genes by genetic variants were only a preliminary exploration of the possible mechanisms, and gene expression quantitative trait loci analysis need to be validated in the future studies. Last, our findings are based on European cohort make it difficult to represent the universal conclusions for other ethnic groups.

In conclusion, our study showed that AD may causally affect severe COVID-19, which were of clinical significance for particular attention to individuals with AD during this pandemic. Conversely, severe COVID-19 may induce AD, indicating that the future study should pay attention to the prognosis of COVID-19.

## Availability of data and materials

All data generated or analyzed during this study are included in this published article and its supplementary information files.

## Data Availability

https://www.pasteur-lille.fr

https://www.covid19hg.org/results/

---

**Figure S1.**
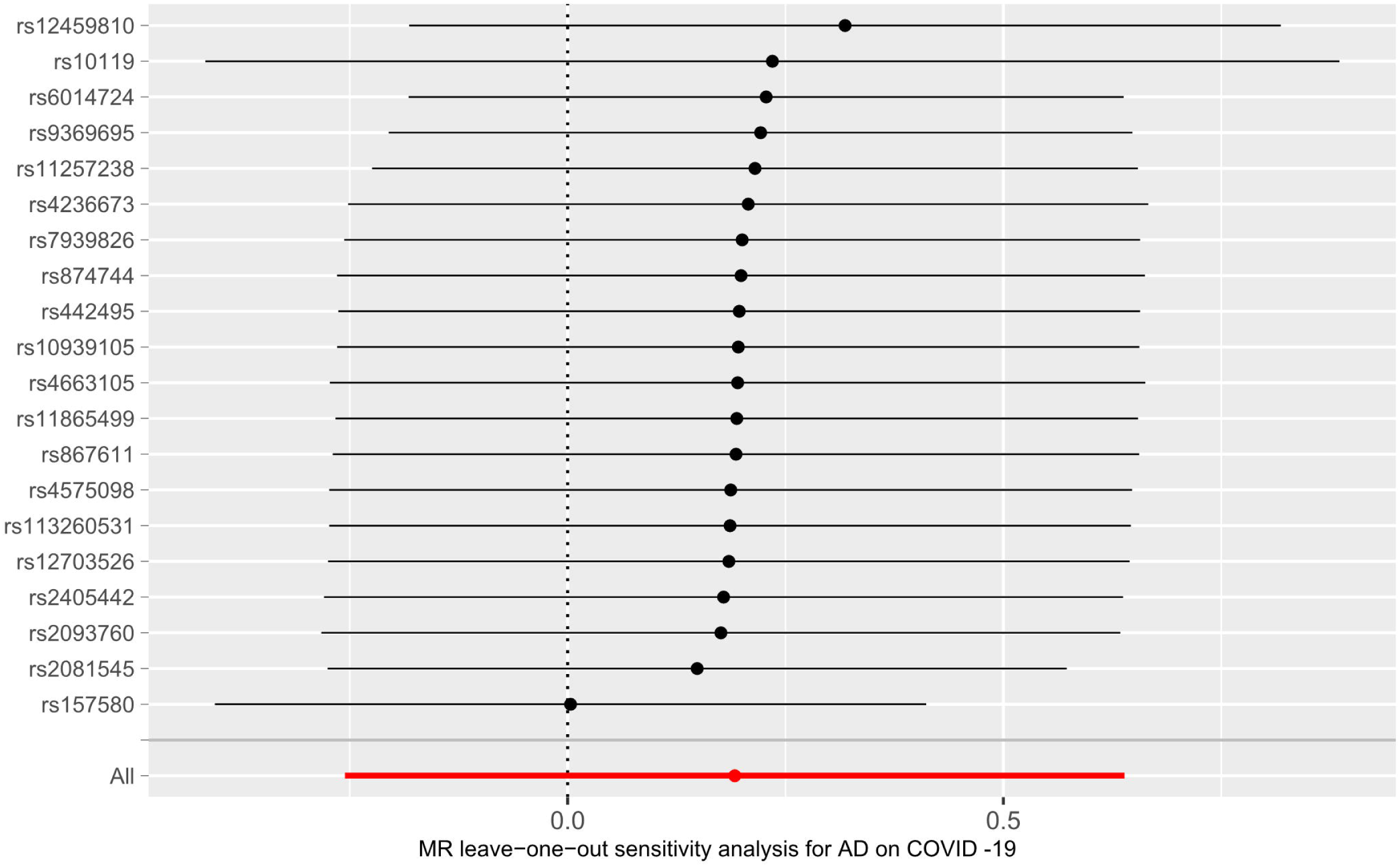
Leave-one-out analysis: each row represents a MR analysis of AD on COVID-19 using all instruments expect for the SNP listed on the y-axis. The point represents the odds ratio with that SNP removed and the line represents 95% confidence interval. AD: Alzheimer’s disease; COVID-19: Coronavirus disease 2019; MR: mendelian randomization; SNP, single-nucleotide polymorphism

**Figure S2.**
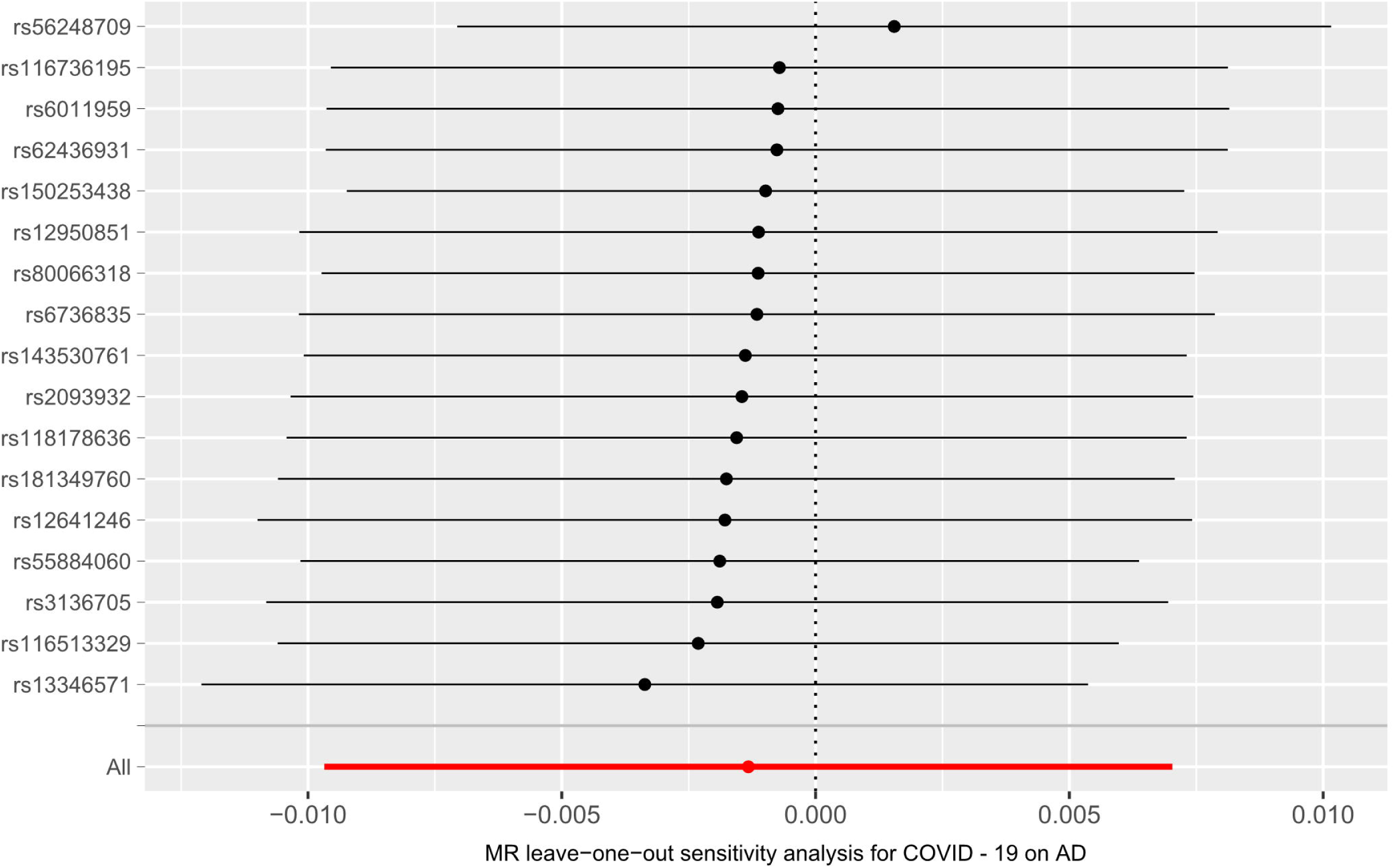
Leave-one-out analysis: each row represents a MR analysis of COVID-19 on AD using all instruments expect for the SNP listed on the y-axis. The point represents the odds ratio with that SNP removed and the line represents 95% confidence interval. AD: Alzheimer’s disease; COVID-19: Coronavirus disease 2019; MR: mendelian randomization; SNP, single-nucleotide polymorphism

